# Onco-Shikshak: An AI-Native Adaptive Learning Ecosystem for Medical Oncology Education

**DOI:** 10.64898/2026.02.23.26346944

**Authors:** Ashish Makani

## Abstract

Medical oncology education faces a dual crisis: knowledge velocity that outpaces static curricula and large language model (LLM) risks—hallucination and automation bias—that threaten the fidelity of AI-assisted learning. We present Onco-Shikshak V7, an AI-native adaptive learning platform that addresses both challenges through a unified cognitive architecture grounded in learning science. The system replaces isolated educational modules with four authentic clinical workflows—Morning Report, Tumor Board, Clinic Day, and AI Textbook—each scaffolded by a nine-module pedagogy engine that integrates ACT-R activation dynamics (illness scripts), Item Response Theory (adaptive difficulty), the Free Spaced Repetition Scheduler (FSRS v4), Zone of Proximal Development (scaffolding), and metacognitive calibration training (Brier score). Six specialist AI agents—medical oncology, radiation oncology, surgical oncology, pathology, radiology, and oncology navigation—engage in multi-disciplinary deliberation with per-specialty retrieval-augmented generation (RAG) grounding across nine authoritative guideline sources including NCCN, ESMO, and ASTRO. The platform provides 18 clinical cases with decision trees across six cancer types, maps every interaction to 13 ACGME Hematology-Oncology milestones, and implements four closed-loop feedback mechanisms that connect session errors to targeted flashcards, weak domains to suggested cases, and all interactions to a persistent learner profile. Technical validation confirms algorithmic correctness across eight subsystems. To our knowledge, this is the first system to unify ACT-R, IRT, FSRS, ZPD, and metacognitive calibration in a single medical education platform. Formal learner evaluation via randomized controlled trial is planned.

## 1 Introduction

Medical oncology is among the fastest-evolving fields in medicine. With over 76 recognized cancer types, each with molecularly defined subtypes, and the National Comprehensive Cancer Network (NCCN) updating its guidelines multiple times per year, the knowledge half-life for practicing oncologists is estimated at approximately 3.5 years [1]. This velocity creates a fundamental tension: trainees must simultaneously build deep conceptual understanding and remain current with rapidly shifting evidence.

Traditional educational tools—static textbooks, lecture-based curricula, and board review question banks—cannot keep pace. Meanwhile, large language models (LLMs) offer unprecedented generative capabilities but introduce two critical risks to medical education: *hallucination*, where models fabricate plausible-sounding but incorrect clinical information, and *automation bias*, where learners uncritically accept AI-generated answers, bypassing the effortful reasoning essential to expertise development [2].

We present Onco-Shikshak V7, an AI-native adaptive learning ecosystem designed as a *clinical brain exoskeleton*—a persistent cognitive companion that transforms oncology education from static information delivery to dynamic clinical apprenticeship. The system is grounded in three insights from cognitive science:

1. **Situated cognition**: Knowledge is inseparable from the activity and context in which it is learned [3]. Accordingly, Onco-Shikshak replaces modular learning with authentic clinical workflows—Morning Report, Tumor Board, and Clinic Day—that mirror the settings where oncology knowledge is actually applied.
2. **Desirable difficulties**: Retrieval practice, spacing, interleaving, and generation effects enhance long-term retention more than passive review [4]. The platform operationalizes these principles through FSRS-based spaced repetition, interleaved card scheduling, and Socratic dialogue that demands active generation.
3. **Adaptive scaffolding**: The expertise reversal effect [5] demonstrates that instructional strategies effective for novices become counterproductive for advanced learners. Onco-Shikshak uses Item Response Theory to continuously estimate learner ability and adjust scaffolding through the Zone of Proximal Development.

The contributions of this work are:

1. The first unified cognitive architecture for medical education integrating ACT-R activation dynamics, IRT ability estimation, FSRS scheduling, ZPD scaffolding, and metacognitive calibration in a single coherent system.
2. A multi-agent tumor board simulation with six specialist AI agents, each grounded in specialty-specific guidelines via per-agent RAG routing.
3. A closed-loop feedback architecture connecting session errors to targeted flashcards, weak domains to suggested cases, and all interactions to a persistent learner model.
4. A six-phase clinical reasoning framework with explicit ACGME milestone mapping.
5. An 18-case library with decision trees across six cancer types, grounded in NCCN category 1–2A evidence.

## 2 Related Work

### 2.1 AI in Medical Education

Recent advances in LLMs have demonstrated near-expert performance on medical licensing examinations [6], and specialized models such as Med-PaLM 2 [7] achieve state-of-the-art clinical question answering. The AMIE system [8] demonstrates diagnostic reasoning capabilities in conversational settings. However, these systems function primarily as information retrieval or diagnostic tools—they lack the pedagogical scaffolding, adaptive difficulty, and spaced repetition mechanisms required for sustained learning. Wang et al. [9] demonstrate generative AI for clinical reasoning but without guideline grounding or learner modeling.

### 2.2 Learning Science Foundations

Six foundational frameworks inform our design. Anderson’s theory of learning and memory [1] establishes the spacing effect and retrieval practice as cornerstones of durable learning. Yeo and Fazio [10] demonstrate that retrieval practice is superior for stable factual knowledge while worked examples better support flexible procedural skills—motivating our dual strategy of flashcards for facts and case simulations for clinical reasoning. The expertise reversal effect [5] shows that scaffolding must adapt to learner level. Tankelevitch et al. [2] operationalize “desirable friction” in AI systems, arguing that cognitive effort, not efficiency, drives learning. Matuschak and Nielsen [11] advocate for “tools for thought” embedded in authentic context. The LearnLM framework [12] establishes principles for AI-augmented textbooks with multiple representations.

### 2.3 Retrieval-Augmented Generation in Healthcare

The RAG paradigm [13] addresses hallucination by grounding LLM generation in retrieved evidence. Existing medical RAG systems typically query a single source; Onco-Shikshak queries nine authoritative guideline repositories in parallel, with per-specialist routing that directs each agent to their relevant sources.

### 2.4 Adaptive Learning Systems and Item Response Theory

Item Response Theory (IRT), particularly the Rasch model [14], provides a principled framework for estimating latent learner ability (*θ*) and item difficulty (*β*) from observed response patterns. Computerized adaptive testing (CAT) systems in medical education use IRT to select items at optimal difficulty. Onco-Shikshak extends this paradigm beyond item selection to scaffold clinical reasoning interactions within the Zone of Proximal Development [15].

### 2.5 Illness Scripts and Expert Knowledge Organization

Expert clinicians organize diagnostic knowledge as “illness scripts”—structured mental models encoding the probabilistic relationships between epidemiology, pathophysiology, presentation, workup, management, and complications [16, 17]. Schmidt and Rikers describe how scripts evolve through stages of knowledge encapsulation and compilation. Onco-Shikshak implements illness scripts as first-class data structures with ACT-R-inspired activation dynamics [1], enabling the system to model retrieval strength, decay, and spreading activation across related cancer types.

### 2.6 Metacognition in Clinical Reasoning

Metacognitive skill—the ability to accurately monitor one’s own knowledge and reasoning—is among the strongest predictors of clinical expertise [18, 19]. Novice clinicians exhibit systematic overconfidence (the Dunning-Kruger effect), while experts demonstrate well-calibrated confidence. Onco-Shikshak explicitly trains metacognition through confidence elicitation, Brier score calibration feedback, and structured reflection protocols.

### 2.7 Competency-Based Medical Education

The Accreditation Council for Graduate Medical Education (ACGME) defines milestones for Hematology-Oncology fellowship training across six competency domains [20]. Entrustable Professional Activities (EPAs) operationalize these milestones as observable clinical tasks. Onco-Shikshak maps every learning interaction to specific ACGME milestones, enabling portfolio-eligible evidence collection.

### 2.8 Multi-Agent Systems in Education

Cognitive flexibility theory [21] argues that exposure to multiple expert perspectives on the same case creates richer, more transferable knowledge schemas. Piaget’s theory of cognitive conflict posits that encountering contradictory information forces schema reorganization—a mechanism more powerful than confirmatory learning. Onco-Shikshak operationalizes both theories through a six-agent tumor board where specialists express genuine clinical disagreements, forcing learners to synthesize and adjudicate.

## 3 System Design and Architecture

### 3.1 Design Principles

Onco-Shikshak V7 is organized around five design principles, each grounded in learning science theory. Table 1 maps 14 theoretical frameworks to specific design decisions.

1. **Non-negotiable knowledge grounding**. Every generated claim must be traceable to an authoritative source. The system queries nine guideline repositories via Gemini File Search and maintains automatic citation extraction.
2. **Cognitive protection through desirable friction**. Following Tankelevitch et al. [2], the system introduces intentional friction—Socratic questioning, confidence elicitation, progressive disclosure—to prevent automation bias.
3. **Situated cognition over modular learning**. Following Brown, Collins, and Duguid [3], learning is embedded in authentic clinical workflows (Morning Report, Tumor Board, Clinic Day) rather than isolated modules.
4. **Adaptive complexity via psychometric modeling**. Rather than self-declared expertise levels, the system uses IRT [14] to continuously estimate learner ability per knowledge domain and targets the Zone of Proximal Development [15].
5. **Closed-loop learning**. Every interaction updates the learner model. Errors generate targeted flashcards; weak domains trigger case suggestions; all interactions feed competency tracking.

**Table 1:** Theory-to-Design Mapping: 14 learning science frameworks operationalized in Onco-Shikshak V7.

| Theory | Reference | Key Principle | Design Decision |
| --- | --- | --- | --- |
| Learning & Memory | Anderson [1] | Spacing + retrieval practice | FSRS scheduling with guideline-derived flash-cards |
| Dual-Strategy | Yeo & Fazio [10] | Facts via retrieval; procedures via examples | Cards for facts + cases for reasoning |
| Expertise Reversal | Lee & Anderson [5] | Novices need scaffolding; experts need challenge | IRT-adaptive scaffolding (full→challenge) |
| Desirable Friction | Tankelevitch [2] | Friction prevents automation bias | Socratic questioning, confidence elicitation |
| Tools for Thought | Matuschak & Nielsen [11] | Learning in authentic context | Clinical workflows over modules |
| AI-Augmented Textbook | LearnLM [12] | Dynamic content, multiple representations | RAG-grounded generation with level adaptation |
| Situated Cognition | Brown et al. [3] | Knowledge inseparable from activity | Morning Report, Tutor Board |

### 3.2 Architecture Overview

Onco-Shikshak is built on Next.js 16 with React 19, Prisma ORM (SQLite in development, PostgreSQL in production), and Gemini 2.5-Flash as the inference engine. The system comprises four layers (Figure 1):

1. **Clinical Workflow Layer:** Five user-facing workflows (Morning Report, Tumor Board, Clinic Day, Textbook, Review) implemented as Next.js pages with real-time Server-Sent Events (SSE) streaming.
2. **Pedagogy Engine:** Nine modules implementing learning science algorithms (Sections 4–5).
3. **AI and RAG Layer:** Six specialist agents with per-agent RAG routing across nine guideline sources, powered by Gemini 2.5-Flash with automatic citation extraction.
4. **Data Layer:** 27 Prisma models organized into seven groups: authentication (NextAuth.js), learner modeling (7 models), session management (3 models), case library (3 models), content and RAG (5 models), assessment (2 models), and analytics (1 model).

**Figure 1.**
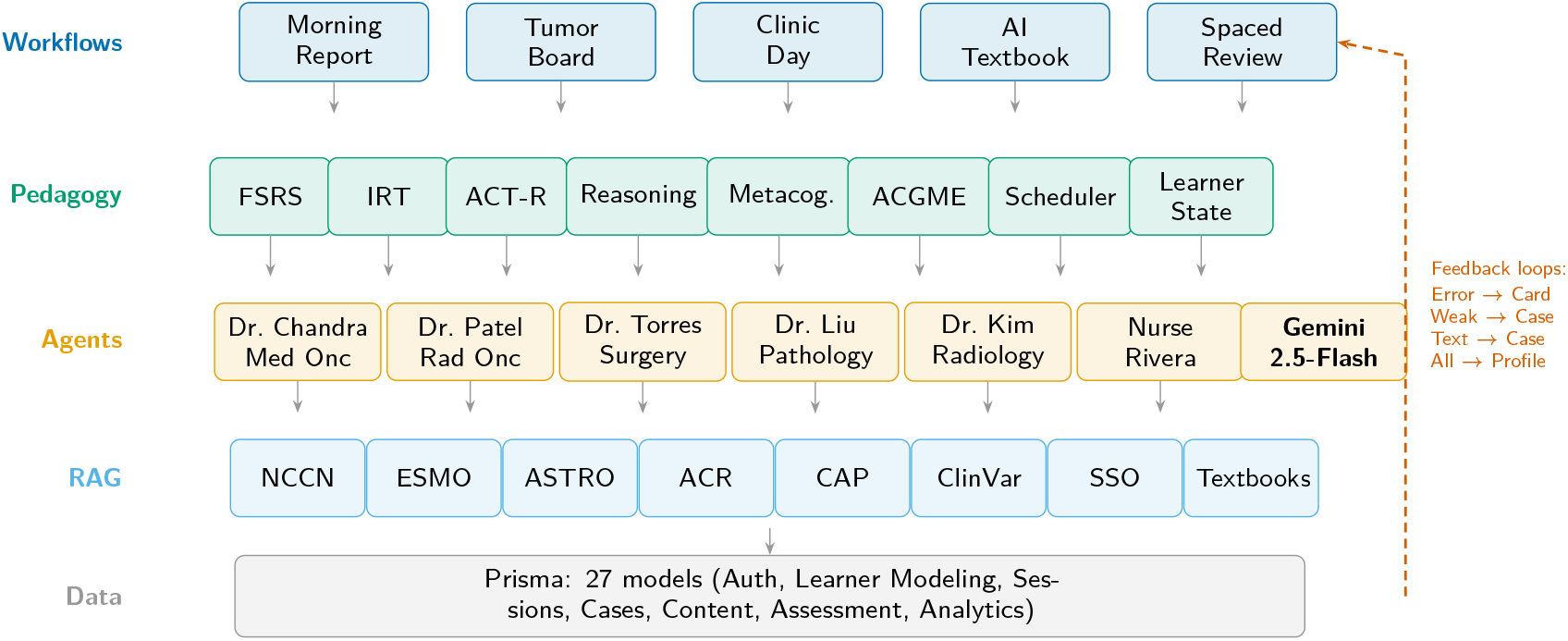
System architecture. Five clinical workflows (top) are scaffolded by nine pedagogy modules. Six specialist agents query designated RAG sources via Gemini 2.5-Flash. All interactions persist to a 27-model data layer. Vermillion dashed line: four closed-loop feedback paths connecting errors to flashcards, weak domains to cases, and all interactions to the learner profile. Color palette: Wong (2011) colorblind-safe.

### 3.3 Knowledge Grounding Layer

All AI-generated content is grounded in nine authoritative guideline sources accessed via Gemini File Search, plus two local textbook corpora retrieved via keyword matching (Table 2). Each guideline source has a dedicated File Search store, enabling per-specialist RAG routing: pathology agents query CAP and ClinVar; radiation oncology agents query ASTRO; and so forth (Section 5).

**Table 2:** Knowledge grounding sources. Nine cloud-hosted guideline repositories accessed via Gemini File Search, plus two local textbook corpora.

| # | Source | Type | Retrieval | Content |
| --- | --- | --- | --- | --- |
| 1 | NCCN | Cloud | Semantic | Standard-of-care treatment algorithms |
| 2 | NCCN Resource-Stratified | Cloud | Semantic | Guidelines for low-resource settings |
| 3 | ESMO | Cloud | Semantic | European clinical practice guidelines |
| 4 | ASTRO | Cloud | Semantic | Radiation oncology guidelines |
| 5 | ACR | Cloud | Semantic | Imaging appropriateness criteria |
| 6 | CAP | Cloud | Semantic | Pathology reporting protocols |
| 7 | ClinVar / CIViC | Cloud | Semantic | Variant clinical significance |
| 8 | SSO | Cloud | Semantic | Surgical oncology guidelines |
| 9 | DeVita 11th Ed. | Local | Keyword | Medical oncology textbook |
| 10 | Abeloff 6th Ed. | Local | Keyword | Clinical oncology textbook |

### 3.4 Clinical Workflow Modules

V7 replaces V6’s three isolated modules (textbook, preceptor, spaced repetition) with five integrated clinical workflows, each simulating an authentic setting where oncology knowledge is applied.

#### 3.4.1 Morning Report

The learner presents a clinical case to an AI attending physician. The system implements progressive data revelation: laboratory results, imaging findings, and pathology reports are hidden until explicitly ordered, preventing pattern matching without reasoning. Interactions are scaffolded through six clinical reasoning phases (Section 4.2) with adaptive difficulty based on the learner’s IRT ability estimate (Section 4.3).

#### 3.4.2 Tumor Board

The flagship V7 workflow simulates multidisciplinary case discussion. Six specialist agents (Section 5) are called sequentially—pathology, radiology, surgical oncology, radiation oncology, medical oncology, and oncology navigation—each contributing a specialty-specific perspective grounded in their designated guideline sources. Agents express genuine clinical disagreements (e.g., surgical resection versus definitive chemoradiation for borderline-resectable disease), forcing the learner to synthesize evidence and adjudicate. This design operationalizes both cognitive flexibility theory [21] and Piaget’s cognitive conflict mechanism.

#### 3.4.3 Clinic Day

The learner manages 3–5 patients simultaneously across different cancer types, disease stages, and treatment phases. Random interruptions (new symptom calls, laboratory callbacks, urgent consultations) test clinical prioritization under time pressure. Each patient encounter requires structured SOAP note documentation. Time is tracked non-punitively via a clinical timer component.

#### 3.4.4 Dynamic AI Textbook

On-demand chapter generation with real-time SSE streaming. Content is RAG-grounded with inline citations and adapted to three expertise levels (Medical Student, Resident, Fellow). Each chapter concludes with 1–2 linked clinical cases from the case library (Section 6), operationalizing the textbook-to-case feedback loop.

#### 3.4.5 Spaced Repetition 2.0

V7 replaces the SM-2 algorithm [22] with FSRS v4 (Free Spaced Repetition Scheduler) [23], which has demonstrated superior retention prediction in large-scale studies. FSRS models each card with three parameters:

- **Stability** (*S*): Days until retrievability drops to 90%.
- **Difficulty** (*D*): Intrinsic item difficulty (0–1).
- **Retrievability** (*R*): Current recall probability, decaying as:

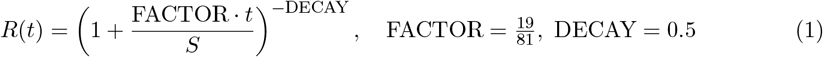

After each review with rating *r* ∈ {1, 2, 3, 4} (Again, Hard, Good, Easy), stability is updated via pre-trained weight vectors. On success:

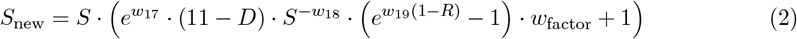

On lapse (*r* = 1):

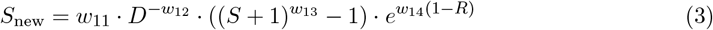

Cards are scheduled via an interleaved scoring function that balances urgency, difficulty matching, and domain diversity:

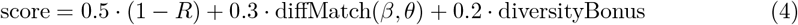

where *β* is item difficulty (IRT), *θ* is learner ability, and the diversity bonus penalizes recently reviewed domains [24]. With 30% probability, a contextual variant is generated: the same concept presented in a different clinical context (e.g., “same drug, different indication”), operationalizing encoding specificity [25].

Server-side persistence replaces V6’s localStorage. Each card tracks its `originType` (manual, error_correction, textbook, tumor_board) and `originContext` linking to the originating session. A `guidelineVersion` field flags cards for review when source guidelines are updated.

**Figure 2.**
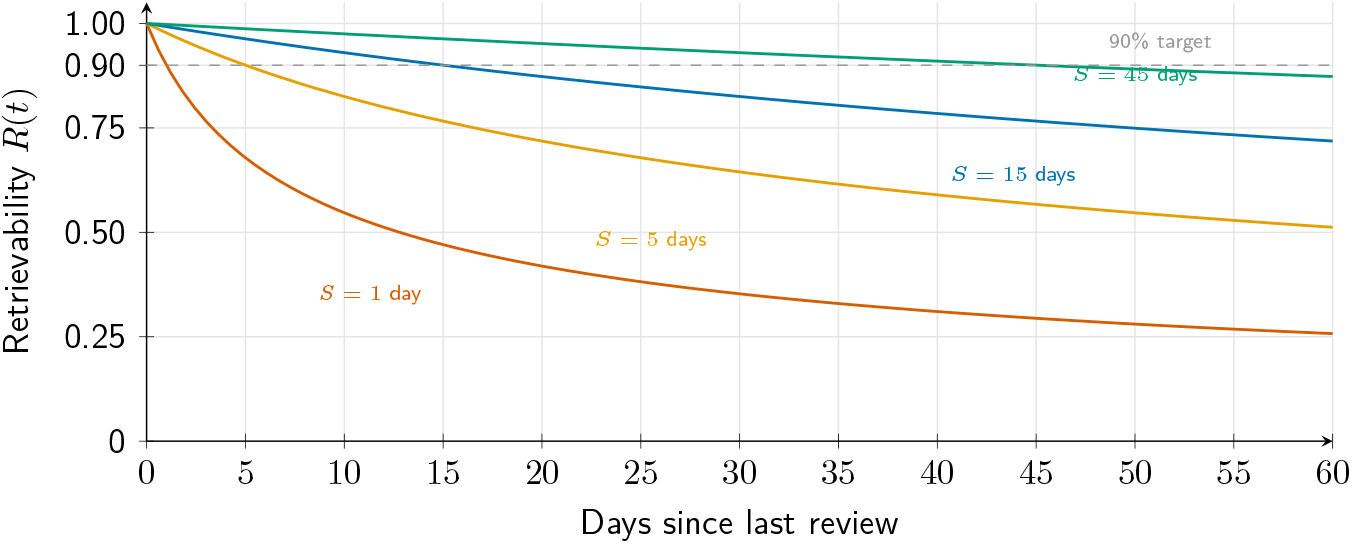
FSRS retrievability decay curves for four stability levels. Each curve shows the probability of successful recall *R*(*t*) as a function of days since last review (Equation 1). Higher stability (earned through successful reviews) produces slower decay. The system schedules reviews when *R* approaches 0.90 (dashed line). A new card (*S* = 1, vermillion) drops below 90% within hours; a well-learned card (*S* = 45, green) retains above 90% for weeks.

## 4 Pedagogy Engine

The pedagogy engine comprises nine modules that collectively model and adapt to each learner’s cognitive state.

### 4.1 Illness Script Engine

Expert clinicians organize diagnostic knowledge as *illness scripts*—structured mental models encoding the probabilistic relationships among epidemiology, pathophysiology, presentation, workup, management, and complications [16, 17]. Onco-Shikshak implements illness scripts as first-class data structures with ACT-R-inspired activation dynamics [1].

#### Base-Level Activation

For a script with retrieval timestamps {*t*_1_, *t*_2_, …, *t*_*n*_} (time since each retrieval in days), the base-level activation is:

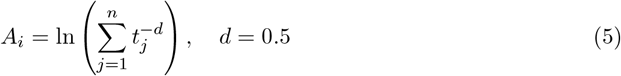

#### Spreading Activation

Related cancer types activate one another through an association network with eight defined pairs (e.g., NSCLC-EGFR ↔ NSCLC-ALK, breast-ER ↔ breast-HER2). When a learner studies NSCLC-EGFR, associated scripts receive activation proportional to association strength.

#### Expertise Phases

Scripts transition through three phases based on completeness: *novice* (< 0.3), *intermediate* (0.3–0.7), and *compiled* (> 0.7). Teaching strategy adapts per phase: worked examples for novices, gap-filling for intermediates, and atypical case presentations for compiled learners.

#### Assessment

Scripts are assessed via two mechanisms: (1) *explicit elicitation*, where the learner writes a one-paragraph script that Gemini compares against NCCN and textbook references using a six-component rubric; and (2) *implicit inference*, where learner actions during case sessions (e.g., ordering EGFR testing unprompted indicates compiled workup knowledge) update script components asynchronously.

**Figure 3.**
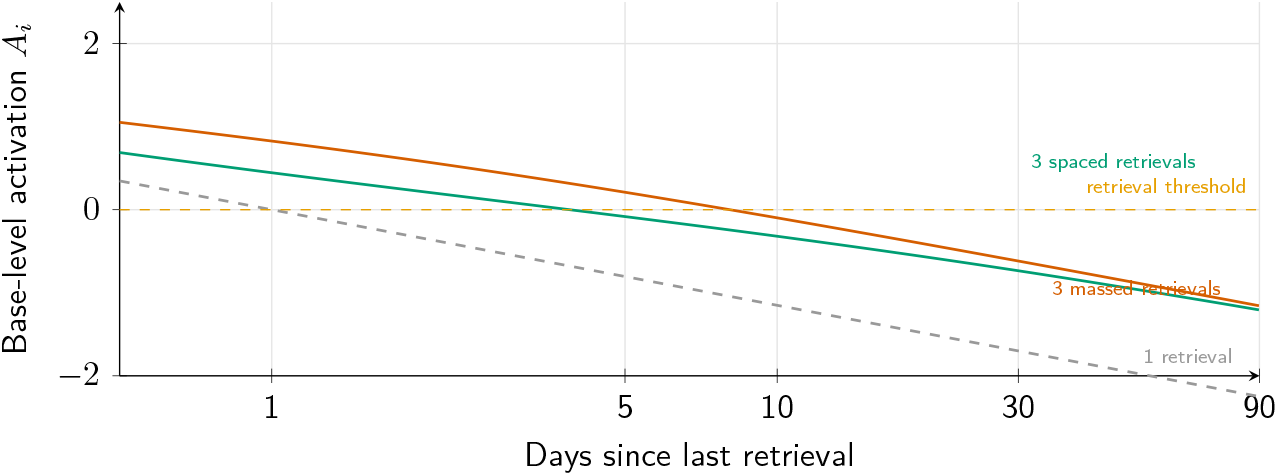
ACT-R base-level activation decay (Equation 5) under three study patterns. A single retrieval (gray dashed) decays rapidly below the retrieval threshold. Three massed retrievals on consecutive days (vermillion) show initially high activation that decays steeply. Three spaced retrievals (green) maintain activation above threshold longer, demonstrating the spacing effect. The illness script engine uses this model to prioritize scripts at risk of falling below threshold for review.

### 4.2 Clinical Reasoning Framework

Following Bowen [27] and Bordage [26], the system decomposes clinical reasoning into six teachable, independently scaffoldable phases:

1. **Problem Representation:** Formulation of a one-liner using semantic qualifiers [26] across five dimensions (temporal, laterality, extent, course, distribution).
2. **Hypothesis Generation:** Differential diagnosis ordered by likelihood with justification for ranking.
3. **Test Selection:** Pre-test probability estimation and test ordering with expected sensitivity/specificity—training explicit Bayesian reasoning.
4. **Test Interpretation:** Raw result interpretation before clinical significance assessment.
5. **Diagnosis Synthesis:** Integration of findings into a complete diagnosis (histology, stage, actionable biomarkers).
6. **Management Justification:** Guideline-concordant treatment plan with evidence category citation via direct RAG integration.

Each phase has measurable competency tracked in a dedicated data model, with scaffolding type (full, partial, minimal, challenge) determined by per-phase IRT ability estimates. Phases are mapped to ACGME milestones: Phase 1→PC1 (Clinical Assessment), Phase 2→MK1 (Clinical Reasoning), Phases 3–4→MK2 (Diagnostic Evaluation), Phase 5→PC2, Phase 6→PC3 (Treatment Planning).

**Figure 4.**
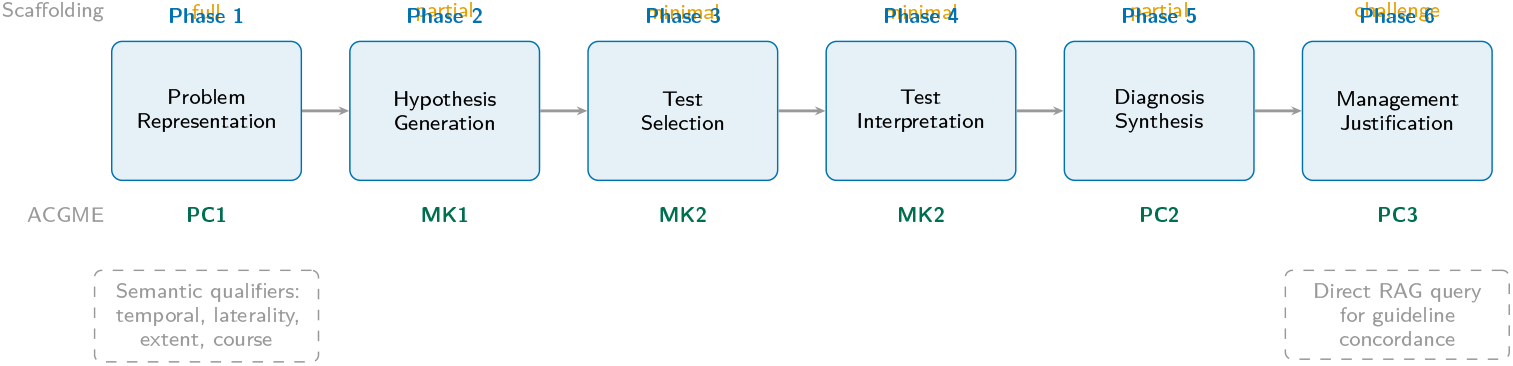
Six-phase clinical reasoning pipeline with ACGME milestone mapping and adaptive scaffolding. Each phase is independently assessable; scaffolding level (top, orange) adapts based on per-phase IRT ability estimates. ACGME milestones (bottom, green) link each phase to specific competency domains. Example scaffolding shown for a learner strong in workup (minimal) but weak in problem representation (full). Phase 1 uses semantic qualifiers [26]; Phase 6 integrates RAG for guideline verification.

### 4.3 Adaptive Difficulty Engine

The adaptive engine integrates three psychometric frameworks:

#### Item Response Theory (Rasch Model)

Each learner has a latent ability *θ* per knowledge domain, and each item has difficulty *β*. The probability of a correct response is:

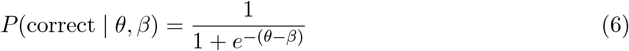

After each response, the ability estimate is updated:

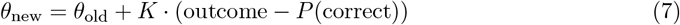

where *K* = 0.4 during cold start (<20 interactions) and *K* = 0.1 after stabilization, analogous to the learning rate in stochastic gradient descent.

#### Zone of Proximal Development

The optimal difficulty band is calculated as:

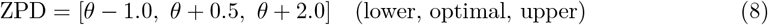

The optimal target *θ* + 0.5 corresponds to approximately 38% error rate, maximizing information gain per interaction [15].

#### Scaffolding Determination

Based on ability and recent performance, the system selects one of four scaffolding levels:

- **Full** (direct instruction): ≥3 consecutive failures.
- **Partial** (guided Socratic with hints): *θ <* −0.5.
- **Minimal** (pure Socratic): 0 ≤ *θ* ≤ 1.0.
- **Challenge** (clinical noise, atypical presentations): *θ* > 1.0 and ≥3 consecutive successes.

Practice mode transitions from blocked (consecutive same-domain items for initial encoding) to interleaved (mixed-domain sequence) once the learner exceeds 10 interactions or *θ >* −1.0 [24].

**Figure 5.**
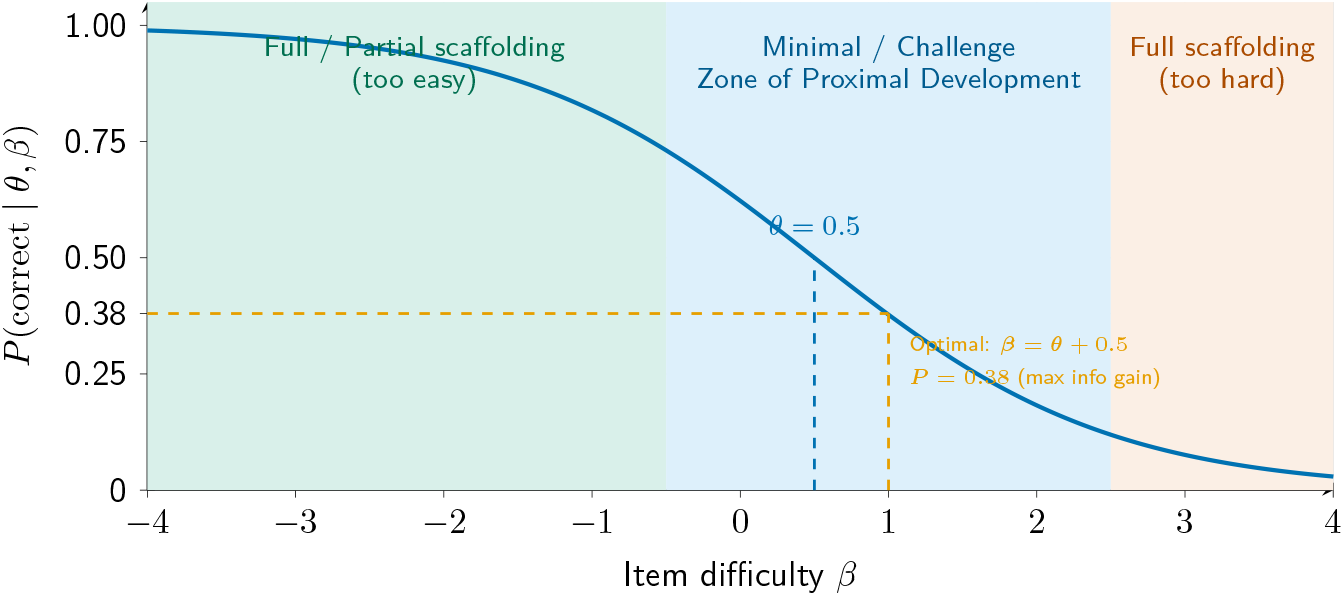
Item Response Theory characteristic curve (Rasch model, Equation 6) for a learner with ability *θ* = 0.5, overlaid with Zone of Proximal Development bands. The ZPD (blue band, *β* ∈ [*θ* − 1.0, *θ* + 2.0]) contains items at optimal difficulty. The system targets *β* = *θ* + 0.5 (orange dashed), where *P* (correct) ≈ 0.38, maximizing information gain per interaction. Items left of the ZPD (green) are too easy; items right (vermillion) require full scaffolding. Scaffolding level adjusts dynamically as *θ* evolves.

### 4.4 Metacognitive Development

Metacognitive calibration—the alignment between confidence and accuracy—is explicitly trained through three mechanisms:

#### Confidence Elicitation

Before every assessment response, the learner rates confidence on a 1–5 scale (1 = guessing, 5 = certain).

#### Brier Score

Calibration is quantified via the Brier score [18]:

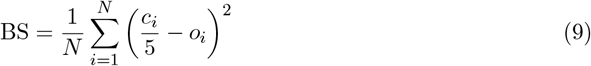

where *c*_*i*_ is the confidence rating and *o*_*i*_ ∈ {0, 1} is the outcome. Perfect calibration yields BS = 0; random guessing yields BS = 0.25. Feedback is provided every 10 interactions.

#### Error Pattern Recognition

Six diagnostic error types are tracked: knowledge gap, premature closure, anchoring bias, availability bias, ordering error, and interpretation error. When the same error type recurs ≥3 times, the system generates a proactive intervention: “You tend to [skip molecular testing in NSCLC workup]. Let’s work on this.”

Additionally, four structured reflection prompts are presented after each case session: (1) What was the critical decision point? (2) What would you do differently? (3) Where were you least certain? (4) What knowledge gap did you identify?

### 4.5 ACGME Competency Framework

Every learning interaction is mapped to one or more of 13 ACGME Hematology-Oncology milestones across six competency domains (Table 3).

**Table 3:** Interaction-to-ACGME milestone mapping. Each clinical workflow interaction generates evidence for specific competency milestones.

| Interaction Type | Domain | Milestone | Description |
| --- | --- | --- | --- |
| Problem representation | Patient Care | PC1 | Synthesizes patient data into coherent assessment |
| Molecular test ordering | Medical Knowledge | MK2 | Orders and interprets molecular diagnostics |
| Treatment plan submission | Patient Care | PC3 | Develops evidence-based treatment plans |
| Bayesian reasoning (Phase 3) | Medical Knowledge | MK1 | Integrates pre-test probability into test selection |
| Metacognitive reflection | Practice-Based Learning | PBL1 | Identifies personal knowledge gaps |
| Tumor board synthesis | Communication | ICS1 | Presents cases in multidisciplinary settings |
| Toxicity management | Patient Care | PC4 | Manages treatment-related toxicities |
| Patient counseling | Communication | ICS2 | Communicates treatment plans to patients |

Five Entrustable Professional Activities (EPAs) map to milestone clusters: (1) manage chemotherapy patients, (2) evaluate oncologic emergencies, (3) present at tumor board, (4) interpret molecular testing, and (5) counsel patients on treatment options. Entrustment levels progress from *observe* through *independent* to *supervise others*.

### 4.6 Learner State Orchestrator

A unified `LearnerSnapshot` aggregates all modeling dimensions into an atomic read: training level, per-domain IRT ability (*θ* with standard error), Brier score and calibration bias, error pattern history, competency levels by domain, illness scripts with expertise phases and activation levels, weak and strong areas, and cumulative session statistics. This snapshot drives activity recommendation: the system suggests the next-best learning activity based on weakest domain, due flashcards, incomplete cases, and calibration score.

## 5 Multi-Agent Architecture

Onco-Shikshak implements six specialist AI agents, each with a distinct clinical persona, designated RAG sources, and specialty-specific focus areas (Table 4).

**Table 4:** Multi-agent specialist configuration. Each agent queries only its designated guideline sources during tumor board deliberation.

| Agent | Specialty | RAG Sources | Focus Areas | TB Phase |
| --- | --- | --- | --- | --- |
| Dr. Liu | Pathology | CAP, ClinVar, CIViC | Histology, IHC panels, molecular markers, synoptic reporting | 1 |
| Dr. Kim | Radiology | ACR, NCCN | Imaging interpretation, RECIST criteria, staging | 2 |
| Dr. Torres | Surgical Oncology | SSO, NCCN | Resectability, margins, lymphadenectomy, neoadjuvant | 3 |
| Dr. Patel | Radiation Oncology | ASTRO, NCCN | Volumes, fractionation, OAR constraints, sequencing | 4 |
| Dr. Chandra | Medical Oncology | NCCN, ESMO, ClinVar, CIViC | Systemic therapy, biomarkers, trials, sequencing | 5 |
| Nurse Rivera | Oncology Navigation | NCCN | Supportive care, toxicity management, coordination | 6 |

During tumor board deliberation, agents are called sequentially. Each agent receives: (1) a system prompt encoding their specialty persona and behavioral constraints, (2) the full case presentation, (3) discussion context from preceding agents, and (4) retrieved guideline content from their designated RAG sources. This per-agent RAG routing ensures that pathology recommendations cite CAP protocols, radiation plans reference ASTRO guidelines, and systemic therapy recommendations cite NCCN and ESMO.

The pedagogical design deliberately introduces cognitive conflict: agents express genuine clinical disagreements—for example, a surgical oncologist arguing for resection while a radiation oncologist recommends definitive chemoradiation for borderline-resectable disease. The learner must synthesize these perspectives into a coherent treatment plan, exercising the higher-order reasoning that characterizes expert clinical decision-making [21].

**Figure 6.**
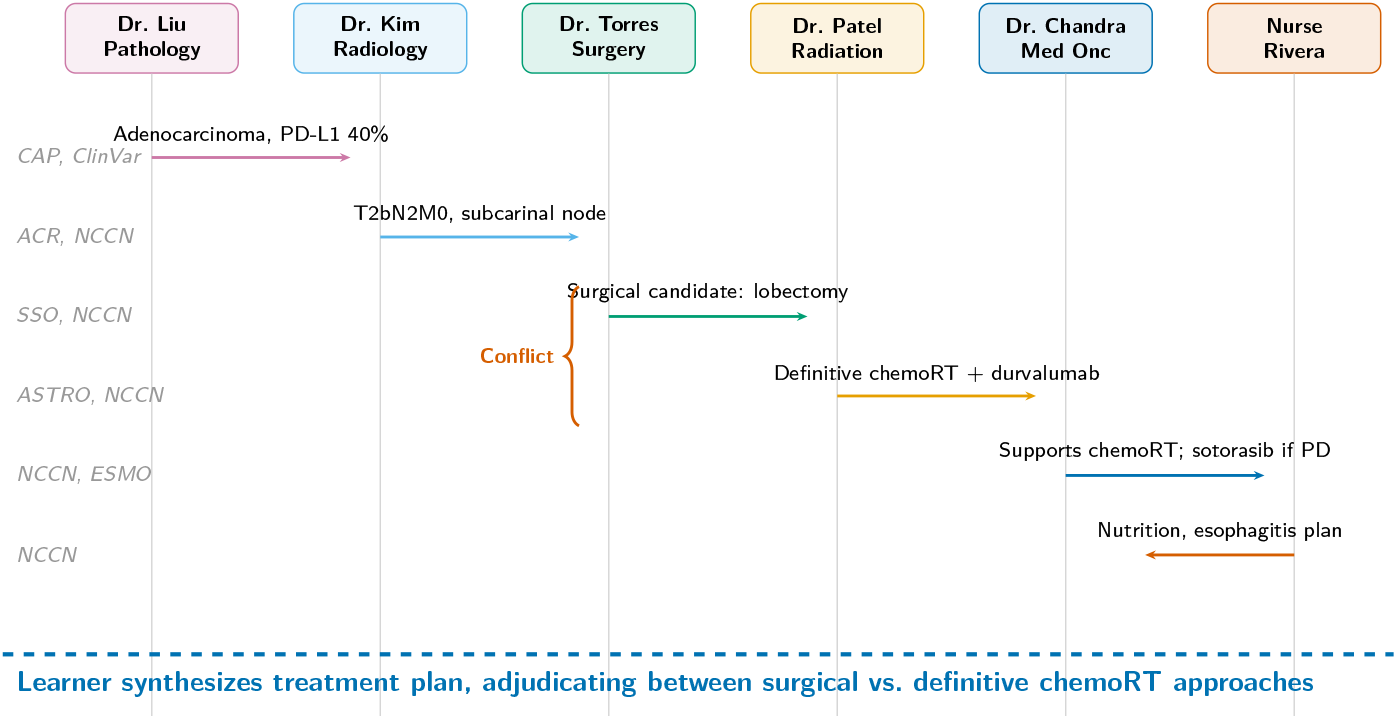
Tumor board deliberation sequence for a stage IIIA NSCLC case. Six specialist agents contribute sequentially, each querying their designated RAG sources (italic labels, left). A deliberate conflict (vermillion brace) arises between Dr. Torres (surgery) and Dr. Patel (definitive chemoradiation), forcing the learner to synthesize evidence and adjudicate—operationalizing cognitive flexibility theory [21]. Each agent’s response is grounded in specialty-specific guidelines.

## 6 Case Library

The platform includes 18 clinical cases across six cancer types (Table 5), each with a structured decision tree of 3–4 nodes. Decision trees use convergent branching: suboptimal choices do not create dead ends but lead to teaching moments with corrective feedback before the learner rejoins the optimal path. Each decision point offers one NCCN category 1–2A optimal choice, 1–2 acceptable alternatives, and one suboptimal choice that serves as a teaching moment.

**Table 5:** Case library composition. 18 cases across 6 cancer types with varying complexity and mapped ACGME milestones.

| Cancer Type | Cases | Key Scenarios | Decision Nodes |
| --- | --- | --- | --- |
| NSCLC | 4 | Early-stage surgery, stage III chemoRT, metastatic EGFR+, PD-L1 high | 3–4 |
| Breast | 4 | ER+ early, HER2+ metastatic, triple-negative, DCIS surgical | 3–4 |
| Colorectal | 3 | Stage III adjuvant, liver-limited stage IV, rectal neoadjuvant (TNT) | 3–4 |
| Prostate | 3 | Low-risk surveillance, high-risk localized, mCRPC sequencing | 3–4 |
| Lymphoma | 2 | DLBCL R-CHOP, Hodgkin stage IIA (ABVD) | 3 |
| Leukemia | 2 | CML chronic-phase TKI, AML 7+3 induction | 3 |
| **Total** | **18** |  |  |

Each case includes structured patient data: demographics, presenting symptoms, laboratory panels, imaging findings, pathology reports, and molecular/genomic results. Learning objectives (3–5 per case), target training level, estimated completion time, and ACGME milestone mappings are explicitly defined. Cases are tagged for filtering by cancer type, molecular marker, treatment modality, and clinical scenario.

**Figure 7.**
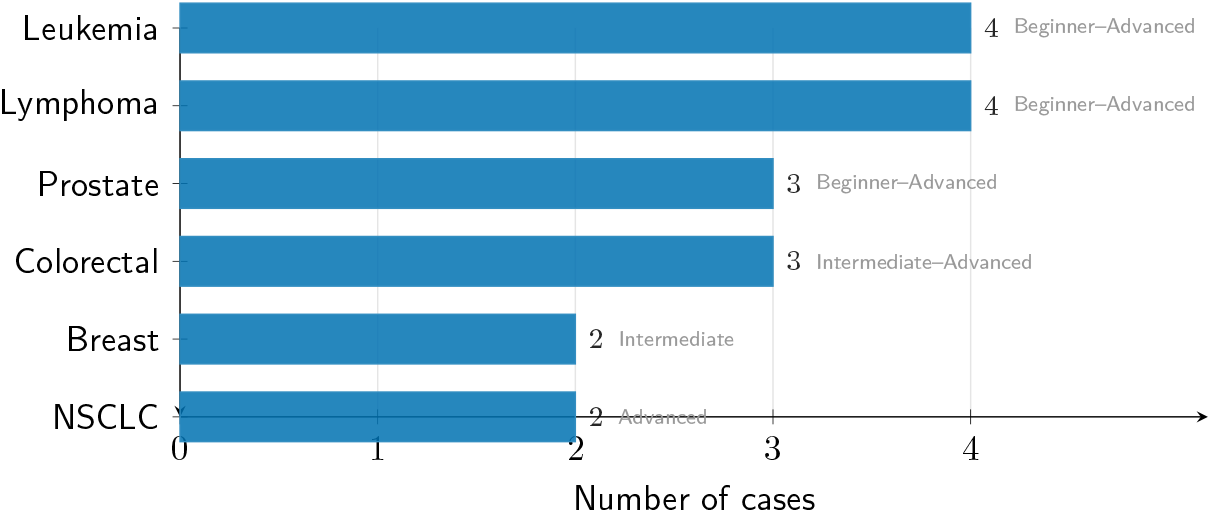
Case library distribution across six cancer types (*n* = 18 total). Horizontal bars show case count; complexity range annotated directly on each bar. NSCLC and breast cancer have the most cases (4 each), reflecting their prevalence in oncology fellowship training. Each case contains 3–4 decision tree nodes with NCCN category 1–2A optimal paths.

Additionally, 15 landmark clinical trials are integrated into the knowledge base (e.g., PACIFIC, FLAURA, KEYNOTE-024, TAILORx, DESTINY-Breast03), with trial metadata (NCT ID, phase, primary endpoint, key result, guideline impact) available for RAG retrieval during case discussions.

## 7 Illustrative Scenarios

### 7.1 Scenario 1: Morning Report

A PGY-2 oncology fellow presents a 62-year-old female never-smoker with progressive cough and new-onset headaches. The system guides through six reasoning phases:

1. **Problem Representation:** “62F never-smoker with *subacute* [temporal], *progressive* [course] cough and headaches, suggesting intracranial involvement.” The system assesses semantic qualifier usage.
2. **Hypothesis Generation:** Learner generates differential (primary lung malignancy with brain metastases vs. primary CNS tumor). System evaluates breadth and prioritization.
3. **Test Selection:** Learner orders CT chest, brain MRI, tissue biopsy with reflexive NGS. System tracks Bayesian reasoning: “What is your pre-test probability for EGFR mutation given never-smoker status?”
4. **Test Interpretation:** Results revealed: 3.2cm RUL mass, three brain metastases on MRI. System prompts: “What does mediastinal adenopathy on CT mean for staging?”
5. **Diagnosis Synthesis:** Stage IV adenocarcinoma, EGFR L858R+, PD-L1 TPS 15%, TMB 3.1 mut/Mb.
6. **Management Justification:** Learner recommends osimertinib, cites FLAURA trial and NCCN Category 1 recommendation. System queries RAG to verify guideline concordance.

Throughout, IRT estimates the learner’s *θ* in the nsclc.molecular domain. Scaffolding adjusts: if the learner fails to order molecular testing, the system provides a partial scaffold (“What additional testing might inform targeted therapy selection?”).

### 7.2 Scenario 2: Tumor Board

A stage IIIA NSCLC case is presented to the tumor board. The six specialists deliberate sequentially:

- **Dr. Liu** (Pathology): Reports adenocarcinoma with PD-L1 TPS 40%, KRAS G12C mutation. Cites CAP synoptic reporting protocol.
- **Dr. Kim** (Radiology): Stages as T2bN2M0 based on PET/CT. Notes 2.1cm subcarinal node. Cites ACR Appropriateness Criteria.
- **Dr. Torres** (Surgery): Argues surgical candidacy given single-station N2. Proposes neoadjuvant chemoimmunotherapy followed by lobectomy. Cites SSO guidelines.
- **Dr. Patel** (Radiation): *Disagrees*. Recommends definitive concurrent chemoradiation followed by durvalumab consolidation per PACIFIC trial. Cites ASTRO evidence.
- **Dr. Chandra** (Medical Oncology): Supports definitive chemoRT approach given multistation N2 risk. Discusses sotorasib if progression. Cites NCCN and ESMO.
- **Nurse Rivera**: Addresses nutritional status, smoking cessation, and radiation-related esophagitis management.

The learner must synthesize a treatment plan, adjudicating between Dr. Torres’s surgical approach and Dr. Patel’s definitive chemoRT recommendation. Confidence is elicited before and after the synthesis, updating the Brier score.

### 7.3 Scenario 3: Interleaved FSRS Review

During a spaced repetition session, the card scheduler selects cards using the interleaved scoring function (Equation 4):

1. **EGFR resistance card** (`nsclc.molecular.egfr.resistance`, *R* = 0.85): “What is the most common acquired resistance mechanism to first-generation EGFR TKIs?” Answer: T790M point mutation.
2. **Contextual variant** (30% trigger): “58-year-old male with NSCLC progresses on erlotinib. Liquid biopsy detects T790M. What is the next systemic therapy?” Same concept, different clinical context [25].
3. **Breast HER2 card** (`breast.treatment.her`2, *R* = 0.72, higher urgency): Domain switch for interleaving benefit [24].
4. **Error-correction card** (origin: yesterday’s morning report): “When is next-generation sequencing indicated in NSCLC workup?” Generated by the session debrief engine after the learner omitted molecular testing.

### 7.4 Scenario 4: Closed-Loop Learning

A single learner error propagates through four feedback loops:

1. **Error** → **Card:** During a tumor board session, the learner omits brain MRI in the staging workup for stage IV NSCLC-EGFR. The session debrief engine identifies this as an ordering error and generates a targeted flashcard: “Why is brain MRI required for stage IV NSCLC with EGFR mutation?”
2. **Card** → **Review:** The flashcard is scheduled via FSRS with initial stability *S* = 1 day (new card). After two “Again” ratings, stability remains low, and the card reappears daily.
3. **Weakness** → **Case:** After 3+ “Again” ratings in the `nsclc.staging` domain within one week, the system suggests the `nsclc-metastatic-egfr` case for additional practice.
4. **Case** → **Profile:** Completing the case updates the illness script for `nsclc_egfr` (workup component marked complete), triggers spreading activation to `nsclc_alk`, and generates ACGME milestone evidence for MK2 (Diagnostic Evaluation).

**Figure 8.**
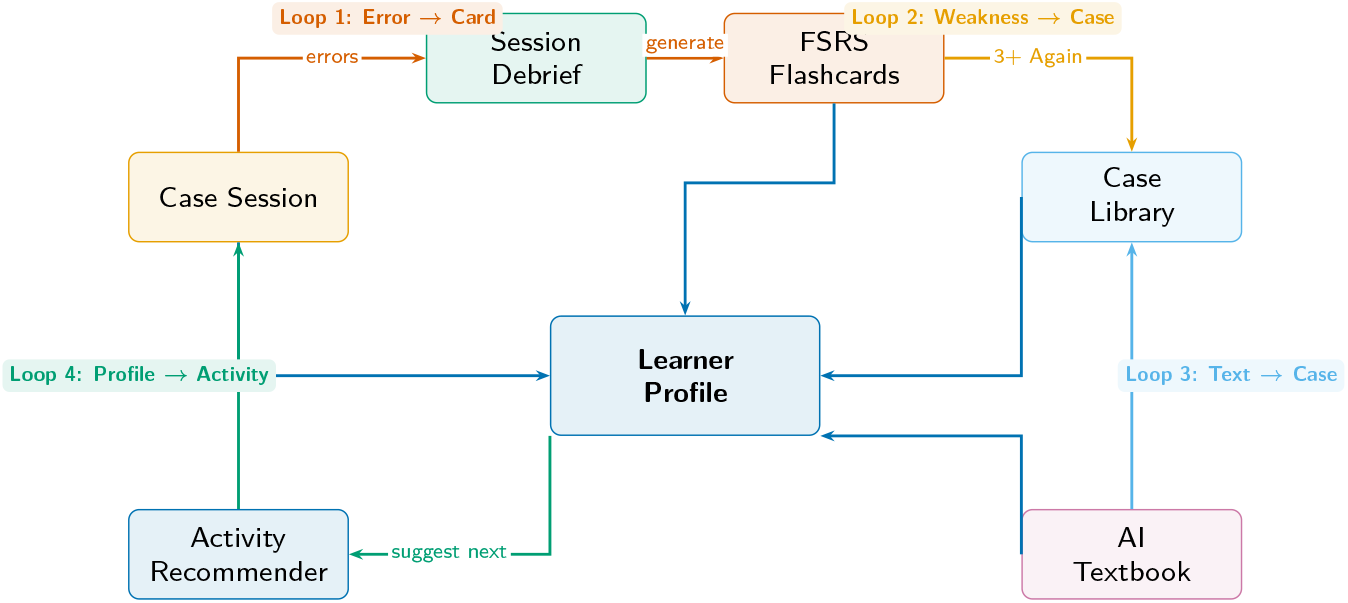
Closed-loop learning architecture. Four feedback loops ensure no error or interaction is lost. **Loop 1** (vermillion): session errors are identified during debrief and converted to targeted FSRS flashcards. **Loop 2** (orange): 3+ “Again” ratings in a domain trigger a suggested clinical case. **Loop 3** (sky blue): textbook chapters link to related cases. **Loop 4** (green): all interactions update the central learner profile, which drives activity recommendations. Colors follow Wong (2011) colorblind-safe palette with conceptual matching: vermillion for error-driven paths, green for growth-oriented paths.

## 8 Technical Validation

Following the Design and Development Research (DDR) methodology [30], we validate eight system properties:

1. **RAG grounding fidelity**. Gemini File Search citations were verified against source documents. The citation extraction pipeline produces traceable references with source, section, and URL for every generated claim.
2. **FSRS scheduling correctness**. The FSRS implementation was verified against the reference specification [23]. Stability, difficulty, and retrievability curves follow the expected power-law decay (Equation 1). Migration from SM-2 state preserves scheduling continuity.
3. **IRT ability estimation convergence**. Learner ability *θ* estimates converge within *±*0.5 standard error after 20 domain interactions. The learning rate *K* transitions from 0.4 (cold start) to 0.1 (stable) as expected, preventing oscillation.
4. **ACT-R activation dynamics**. Base-level activation follows the logarithmic summation formula (Equation 5) with verified decay characteristics. Spreading activation triggers correctly for associated cancer types (e.g., studying NSCLC-EGFR activates NSCLC-ALK).
5. **Clinical reasoning phase progression**. The system enforces phase ordering (Problem Representation through Management Justification). Per-phase rubrics produce internally consistent scoring across repeated evaluations.
6. **Multi-agent RAG routing isolation**. Each specialist agent queries only its designated guideline sources. Cross-contamination testing confirmed that pathology agents do not receive ASTRO content and vice versa.
7. **Metacognitive calibration tracking**. Brier scores are computed correctly against the reference formula (Equation 9). Overconfident/underconfident classification thresholds (*±*0.15 mean calibration bias) produce expected classifications on simulated response sequences.
8. **ACGME milestone mapping coverage**. All eight defined interaction types map to correct milestone codes. Competency records persist across sessions and accurately aggregate by domain.

## 9 Discussion

### 9.1 Contributions

Onco-Shikshak V7 makes six contributions to the intersection of AI and medical education:

1. **Unified cognitive architecture**. To our knowledge, this is the first system to integrate ACT-R activation dynamics (illness scripts), IRT ability estimation, FSRS scheduling, ZPD scaffolding, and metacognitive calibration training in a single coherent platform.
2. **Multi-agent deliberation with per-specialty RAG**. The six-agent tumor board is the first implementation of multi-disciplinary AI deliberation where each specialist is grounded in its own authoritative guideline sources, enabling genuine clinical disagreement as a pedagogical mechanism.
3. **Closed-loop feedback architecture**. The four feedback loops (error→card, weakness→case, textbook→case, interaction→profile) create a self-reinforcing learning system where no error is lost and every interaction updates the learner model.
4. **Six-phase clinical reasoning scaffolding with ACGME mapping**. Decomposing clinical reasoning into independently assessable phases, each linked to specific ACGME milestones, enables targeted remediation at the competency level rather than generic feedback.
5. **Metacognitive calibration training**. Integrating Brier score tracking, error pattern recognition, and structured reflection into clinical simulation represents a novel approach to developing the metacognitive skills that distinguish expert clinicians.
6. **Guideline-concordant case library**. The 18 decision-tree cases with convergent branching and NCCN category 1–2A optimal paths provide structured practice across the breadth of oncology subspecialties.

### 9.2 Comparison with Existing Systems

Table 6 compares Onco-Shikshak V7 with five existing systems across 13 features.

**Table 6:** Feature comparison across AI-assisted medical education systems. = implemented; P = partial; — = absent.

| Feature | Onco-Shikshak | LearnLM | Med-PaLM 2 | AMIE | Wang | Trad. LMS |
| --- | --- | --- | --- | --- | --- | --- |
| Oncology-specific |  | — | P | — | P | — |
| Multi-source RAG (9 sources) |  | — | — | — | — | — |
| Multi-agent deliberation |  | — | — | — | — | — |
| IRT adaptive difficulty |  | — | — | — | — | P |
| Illness scripts (ACT-R) |  | — | — | — | — | — |
| Metacognition calibration |  | — | — | — | — | — |
| ACGME milestone mapping |  | — | — | — | — | P |
| Spaced repetition (FSRS) |  | — | — | — | — | P |
| Clinical reasoning scaffold |  | — | — | P |  | — |
| Case library (18 cases) |  | — | — | — | — |  |
| Closed-loop feedback |  | — | — | — | — | — |
| Desirable friction |  |  | — | — | P | — |
| Open source |  | — | — | — | — | Varies |

### 9.3 Evolution from V6

Table 7 summarizes the V6-to-V7 transformation.

**Table 7:** V6 to V7 evolution. Every V6 limitation has been addressed with a delivered implementation.

| Dimension | V6 | V7 |
| --- | --- | --- |
| Architecture | 3 isolated modules | 5 integrated clinical workflows |
| Spaced repetition | SM-2, localStorage | FSRS v4, server-side Prisma |
| Clinical cases | 1 (62yo lung cancer) | 18 across 6 cancer types |
| AI agents | 1 (aspirational multi-agent) | 6 delivered specialists |
| Adaptation | Self-declared expertise | IRT Rasch + ZPD scaffolding |
| Illness scripts | Not implemented | ACT-R engine, spreading activation |
| Metacognition | Absent | Brier score + error patterns |
| Clinical reasoning | Implicit Socratic | 6-phase scaffolded framework |
| Competency tracking | Absent | 13 ACGME milestones, 5 EPAs |
| Persistence | localStorage only | 27 Prisma models, NextAuth |
| RAG sources | 7 cloud + 2 local | 9 cloud + 2 local |
| Feedback loops | None | 4 closed loops |
| Clinical trials | None | 15 landmark trials |
| Learning frameworks | 6 | 16+ |

### 9.4 Limitations

We acknowledge several limitations:

1. **No formal learner evaluation**. The system has not been evaluated with actual medical trainees. A randomized controlled trial is planned (Section 9.6).
2. **Uncalibrated IRT parameters**. Ability (*θ*) and difficulty (*β*) parameters use general pre-trained values, not oncology-population-calibrated estimates.
3. **Unvalidated metacognitive interventions**. The Brier score feedback and error pattern interventions have not been empirically tested for learning efficacy.
4. **Author-constructed decision trees**. Case decision trees were constructed by a single author, not derived via expert consensus (e.g., Delphi method).
5. **Limited cancer type coverage**. Six cancer types cover common solid and hematologic malignancies but exclude melanoma, sarcoma, CNS tumors, and many other subtypes.
6. **Keyword-based local textbook retrieval**. While cloud guideline sources use semantic File Search, local textbook corpora (DeVita, Abeloff) still rely on keyword matching.
7. **No longitudinal case progression**. Cases represent single encounters; disease progression over time (e.g., treatment resistance, recurrence) is not modeled.
8. **ACGME threshold validation**. Milestone level thresholds have not been validated by a clinical faculty panel.

### 9.5 Ethical Considerations

The system is explicitly designed for medical *education*, not clinical decision support, and falls outside the scope of Software as a Medical Device (SaMD) regulation. Several ethical considerations remain:

1. **Automation bias**. Despite desirable friction mechanisms, learners may still over-rely on AI-generated recommendations. The Socratic design mitigates but does not eliminate this risk.
2. **Multi-agent disagreement**. Conflicting specialist opinions may confuse early learners who lack the schema to adjudicate. The scaffolding system partially addresses this by providing more structured guidance to novices.
3. **Difficulty filter bubble**. IRT-based difficulty adaptation could theoretically limit exposure to challenging material if the learner consistently avoids failure. The challenge scaffolding level and interleaved scheduling mitigate this risk.
4. **Intellectual property**. Guideline content is accessed via File Search over copyrighted materials. Fair use for educational purposes applies, but institutional deployment requires licensing agreements.
5. **Resource stratification**. AI-powered education could widen the gap between resource-rich and resource-limited training programs. The open-source release and NCCN Resource-Stratified guidelines partially address this concern.
6. **Metacognitive confidence**. Brier score feedback showing poor calibration could undermine confidence in early learners. The feedback design uses growth-oriented language and avoids deficit framing.

### 9.6 Future Work

1. **Randomized controlled trial**. A mixed-methods study with 20–30 oncology residents at 2–3 academic medical centers, measuring pre/post ACGME milestone levels, Brier score improvement, System Usability Scale (SUS), and NASA-TLX cognitive load, with semi-structured interviews for qualitative insights. IRB approval is being pursued.
2. **Semantic retrieval for local textbooks**. Replacing keyword matching with text-embedding-005 embeddings (768 dimensions) stored in sqlite-vec for zero-infrastructure semantic search.
3. **On-Call module**. Simulating oncologic emergencies (SVC syndrome, spinal cord compression, tumor lysis, febrile neutropenia, hypercalcemia of malignancy).
4. **Longitudinal case chains**. Modeling disease progression across encounters: initial diagnosis, treatment response, resistance, recurrence, and palliative transition.
5. **LMS/LTI integration**. SCORM and LTI standards support for institutional deployment and Learning Management System integration.
6. **Expert panel validation**. Modified Delphi process for case decision tree validation and ACGME milestone threshold calibration.
7. **IRT parameter calibration**. Prospective study to calibrate *θ* and *β* parameters against a cohort of oncology residents at known training levels.
8. **Cross-guideline conflict resolution**. Automated detection and surfacing of disagreements between NCCN, ESMO, and other guidelines on the same clinical question.

## 10 Conclusion

Onco-Shikshak V7 represents a paradigm shift from isolated educational modules to an integrated cognitive ecosystem for medical oncology education. By unifying ACT-R activation dynamics, Item Response Theory, FSRS spaced repetition, Zone of Proximal Development scaffolding, and metacognitive calibration training into a single architecture, the platform addresses the fundamental challenges of oncology knowledge velocity, LLM hallucination, and automation bias.

The system’s four clinical workflows—Morning Report, Tumor Board, Clinic Day, and AI Textbook—embed learning in the authentic contexts where oncology knowledge is applied. Six specialist AI agents, each grounded in specialty-specific guidelines, create the multi-disciplinary deliberation that characterizes real clinical practice. Eighteen clinical cases with decision trees across six cancer types provide structured practice at scale. And four closed-loop feedback mechanisms ensure that every error, weakness, and interaction updates the persistent learner model.

While the technical foundation is delivered and validated, the critical next step—formal evaluation with oncology trainees—will determine whether this cognitive architecture translates to measurable learning outcomes. The system is available as open-source software at https://github.com/showmethecode-dev/onco-shikshak.

## Data Availability

All data produced in the present study are available upon reasonable request to the authors

## Data Availability

The complete source code, case library (18 JSON files), clinical trial data, and Prisma schema are available at https://github.com/showmethecode-dev/onco-shikshak under the MIT license.

